# High prevalence of asymptomatic malaria in Forest Guinea: Results from a rapid community survey

**DOI:** 10.1101/2023.09.01.23294934

**Authors:** Charlotte C Hammer, Mariama Dalanda Diallo, Boubacar Kann, Fatoumata Sanoh, Tamba N’fantoma Leno, Oumar Mansare, Ismail Diakité, Abdoulaye Djibril Sow, Yacouba Konate, Emilie Ryan-Castillo, Alpha Mahmoud Barry, Claire J Standley

## Abstract

Malaria is endemic in Guinea, however, the extent and role in transmission of asymptomatic malaria are not well understood. In May 2023, we conducted a rapid community survey to determine *Plasmodium falciparum* prevalence among asymptomatic individuals in Middle Guinea (Dalaba) and Forest Guinea (Guéckédou). We used a cluster sampling approach with purposive selection of two prefectures and four communities and full enrolment of all individuals in the selected communities. Prevalence was calculated with a 95% confidence interval (CI). In Dalaba, 6/239 (2.1%, CI 0.9-4.8%) individuals tested positive for P. *falciparum* by rapid diagnostic test, while in Guéckédou, 147/235 (60.9%, CI 54.5-66.9%) of participants tested positive. Asymptomatic malaria needs to be considered more strongly as a driver for transmission when designing control strategies, especially in Forest Guinea and potentially other hyper-endemic settings.

**Key results and their importance:**

1. Prevalence of asymptomatic malaria was very high (60.9%, CI 54.5-66.9%) in the selected communities in Forest Guinea.
2. Prevalence was expectedly low (2.1%, CI 0.9-4.8%) in the selected communities in Middle Guinea.
3. Current control strategies in Forest Guinea seem insufficient to reduce malaria prevalence, and likely also transmission.
4. Both policies and control strategies need to more proactively consider asymptomatic malaria in hyper-endemic settings.

## Introduction

Despite decades of control efforts, malaria remains endemic in many countries in Africa and is a major cause of morbidity and mortality, particularly in children (1). The World Health Organization’s current guidelines focus on “test, treat, and track” strategies of symptomatic individuals, complemented by preventive efforts such as vector control and chemoprophylaxis for high-risk groups (2, 3). However, there is growing recognition that many individuals infected with malaria may not display overt symptoms, due to immune protection from previous exposures and other factors, but yet may still contribute to overall disease transmission (4-6). The extent to which these asymptomatic infections exist in a population may help to determine the effectiveness of existing control efforts. Guinea is highly endemic for malaria, with Forest Guinea being a hyper-endemic area, experiencing year-round transmission (7, 8), while the holo-endemic coastal and middle regions of the country see more seasonal transmission (8-10). Overall malaria incidence ranged from 87-101 per 1000 per year in between 2006-2010 (9). Regional differences in incidence of non-severe malaria ranged from 57-103 per 1000 per year in 2011 (11). In 2021, malaria prevalence among symptomatic and asymptomatic children of 6-59 months ranged from 1.5% (Conakry capital region) to 55.0% (N’Zérékoré, Forest Guinea) (12), and in 2011, parasite prevalence in all age groups ranged from 3% (Conakry capital region) to 66% (Forest Guinea).(11) Since these surveys were conducted, additional control efforts have been implemented in hyper- and holo-endemic settings, notably roll-out of seasonal chemoprophylaxis to high-risk groups. The main parasite present in Guinea is *Plasmodium falciparum* (8).

Given the relative lack of recent data regarding current prevalence of asymptomatic malaria in individuals over the age of five, and subsequentially the potential contribution to transmission, we sought to estimate the prevalence of asymptomatic malaria in older children and adults in Middle Guinea and Forest Guinea.

## Methods and materials

As part of a prospective, longitudinal study on acute febrile illness (AFI) in two prefectures in Guinea, we conducted a rapid community survey in May-June 2023, which is the beginning of the rainy season. Our two study areas were Dalaba, in Middle Guinea and Guéckédou, in Forest Guinea. We used a three-level cluster sampling approach. The two prefectures were chosen purposively as representing the diversity of prevalence and incidence of AFI in Guinea based on past surveillance data. Within each prefecture two communities were chosen purposively based on guidance from the the local health authorities and experts for their suitability, which included consideration of accessibility and research fatigue on the part of the communities. All individuals within these communities were eligible to be enrolled in the study if they were six years old or older, did not currently show any clinical signs of AFI and were able and willing to provide consent either themselves or via their parents/guardians for a socio-demographic and behavioural questionnaire and finger-prick blood sample for a malaria rapid diagnostic test (RDT) to detect *Plasmodium falciparum* using the Bioline™ Malaria Ag P.f test kit from Abbott. Prevalence was calculated with a 95% confidence interval (CI). Data were analysed in R using RStudio.

## Results

In Guéckédou, 239 individuals met the inclusion criteria and in Dalaba, 240. All of them were included in the study with RDT results being available for 474 across both sites. In Guéckédou, 51 % of participants were male and in Dalaba 68 % of participants were male. The median age in in Guéckédou was 20 years (SD 18.54) and in Dalaba it was 30.5 (SD 17.37) years. Figure 1 gives an overview of the age and sex breakdown in both study sites. In Dalaba, 6/239 (2.1%, CI 0.9-4.8%) individuals tested positive for P. *falciparum* by RDT while in Guéckédou, 147/235 (60.9%, CI 54.5-66.9%) of participants tested positive. Positivity decreased with increasing age in both prefectures (see table 1) with children being most affected.

**Table 1:** Prevalence of asymptomatic malaria in the prefectures Dalaba and Guéckédou in May/June 2023 by age group.

| Age | Prevalence Dalaba (95% CI) | Prevalence Gueckedou (95% CI) |
| --- | --- | --- |
| All | 2.09 (0.90-4.80) | 60.85 (54.48-66.87) |
| 6-10 | 6.25 (0.32-28.33) | 82.50 (68.05-91.25) |
| 11-15 | 12.50 (3.50-63.02) | 87.50 (73.89-94.54) |
| 16-20 | 0.00 (0.00-10.15) | 75.61 (60.66-86.17) |
| 21-25 | 3.85 (0.20-18.89) | 46.67 (30.23-63.86) |
| 26-30 | 3.57 (0.18-17.71) | 45.45 (26.92-63.34) |
| 31-35 | 0.00 (0.00-14.87) | 37.50 (13.68-69.43) |
| 36-40 | 0.00 (0.00-12.46) | 42.86 (15.82-74.95) |
| 41-45 | 0.00 (0.00-16.11) | 62.5 (30.57-86.32) |
| 46-50 | 0.00 (0.00-29.91) | 50.00 (21.52-78.48) |
| 51-55 | 0.00 (0.00-25.88) | 16.67 (0.85-56.35) |
| 56-60 | 0.00 (0.00-25.88) | 16.67 (0.85-56.35) |
| 61-65 | 0.00 (0.00-32.44) | 0.00 (0.00-35.43) |
| 66-70 | 0.00 (0.00-48.99) | 16.67 (0.85-56.35) |
| 71-75 | 0.00 (0.00-48.99) | 0.00 (0.00-56.15) |
| 76-80 | 0.00 (0.00-94.87) | 100.00 (5.13-100.00) |
| 81-85 | NaN | NaN |
| 86-90 | 0.00 (0.00-65.76) | 50.00 (2.56-97.44) |
| > 90 | NaN | NaN |

**Figure 1:**
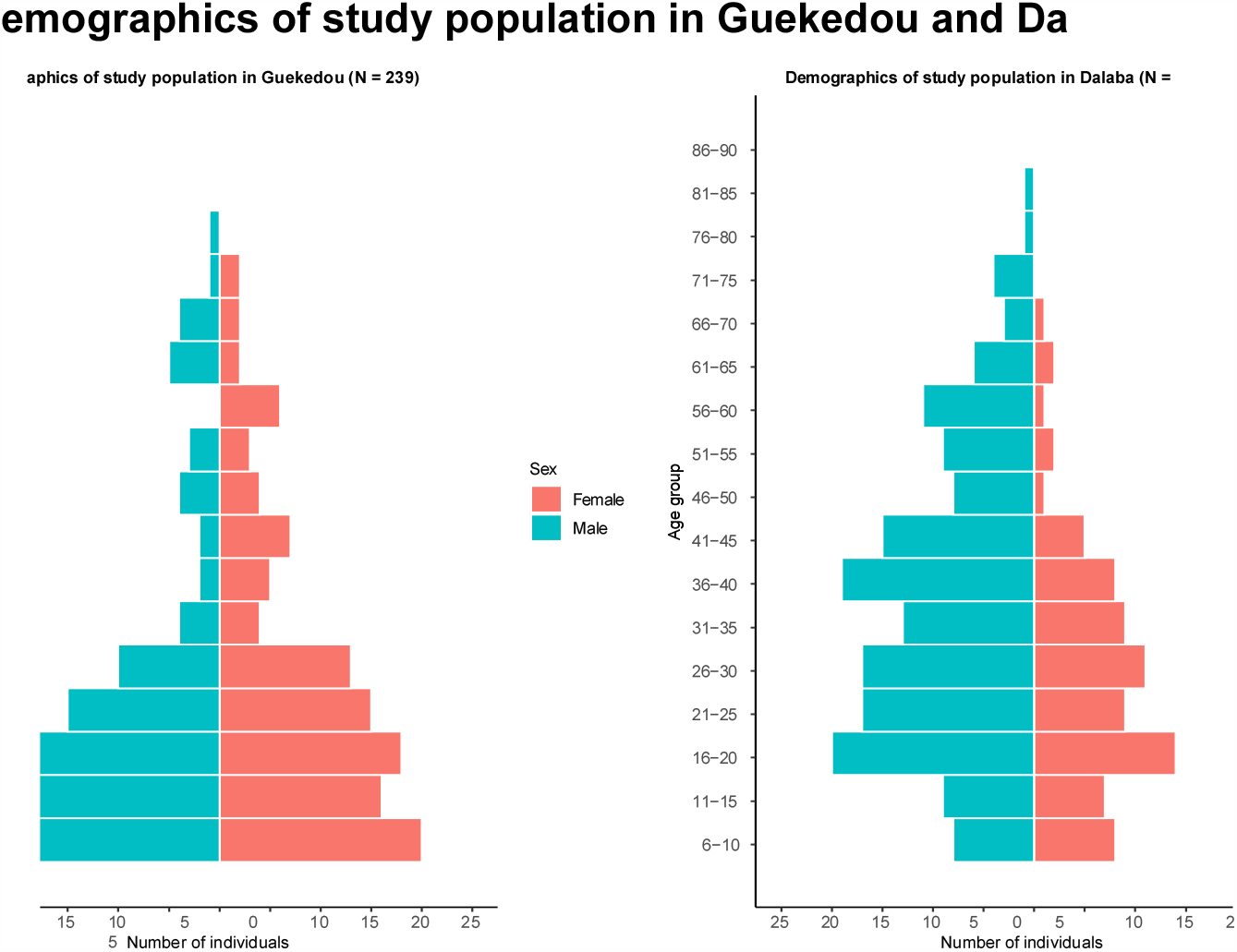
Demographic characteristics of study participants by prefecture. Participants aged below 6 were not eligible for inclusion.

## Discussion and Conclusion

Forest Guinea is a hyper-endemic area for malaria, despite this, the results were still higher than expected given that only asymptomatic individuals were eligible to participate. Results would not have been expected to be as low as in Dalaba which is not in a hyper-endemic zone and in our sample had a higher median age. However, the difference is large enough to not be fully explained by these two factors alone.

The more expected results from Dalaba using the same RDT and eligibility criteria should reduce the likelihood of the results in Guéckédou having been influenced by quality issues with the RDT. The results from the second study side can therefore be seen as a control for the validity of the test kits. The study is potentially biased by the purposive sampling applied when selecting the communities as one of the criteria applied was the possibility for the study team to access the field site, meaning those communities with better road access were more likely to be chosen. Additionally, we only conducted malaria RDTs and not microscopy in this study. For confirmation and further investigation of the results, follow-up with microscopy would be recommended, especially to ascertain risk of transmission through observation of gametocytes.

As Forest Guinea experiences year-round transmission, we expect these results to not be a singular phenomenon, however, we strongly suggest a repeat sampling during or at the end of the rainy season. Dalaba experiences seasonal transmission patterns, with peak incidence between July and October and the beginning of the rainy season seeing lower malaria prevalence (8). We expect the prevalence of asymptomatic malaria to be significantly higher during or at the end of the rainy season on Dalaba.

While additional research is needed to explore these results further, they suggest that current control strategies in Forest Guinea are likely insufficient with respect to malaria transmission, putting children and other vulnerable groups at risk of severe disease. National decision-makers should consider the role of asymptomatic malaria as a driver for transmission in developing policies and intervention strategies in the future. The results also highlight the limitations of test to treat approaches for symptomatic individuals as a disease control strategy as they exclude a significant group of individuals. Further approaches to target and limit transmission in asymptomatic groups may be warranted, in addition to other standard control measures such as vector control and (seasonal) chemoprophylaxis for high-risk groups.

Additionally, we recommend that data collection and analysis for malaria control should also consider older children and adults. Currently, the focus is primarily on children below the age of six. While this is an important risk group for morbidity management, from the point of transmission including other age groups would also be valuable, and help to determine prevalence profiles in other vulnerable groups, such as pregnant women. Ideally, such data should be not only collected regularly but made publicly available for analysis, within the bounds of privacy considerations.

## Data Availability

Due to the very small number of respondents in some age and gender (other) categories, the data cannot be made publicly available as they are potentially identifying.

## Acknowledgements

We would like to thank all participants for their time and availability. We would also like to acknowledge the input from the wider study team.

## Ethics

The study has been approved by the institutional review board of Georgetown University (STUDY00002481) and by the Comité National D’Éthique Pour la Recherche en Santé (CNERS) of the Republic of Guinea (040/CNERS/2022).

## References

1. World Health Organization. World Malaria Report 2022. Geneva: World Health Organization; 2022.

2. World Health Organization. Who Guidelines for Malaria. Geneva: World Health Organization; 2023.

3. World Health Organization. Test. Treat. Track. Scaling up Diagnostic Testing, Treatment and Surveillance for Malaria. Geneva: World Health Organization; 2012.

4. Ibrahim AO, Bello IS, Ajetunmobi AO, Ayodapo A, Afolabi BA, Adeniyi MA. Prevalence of Asymptomatic Malaria Infection by Microscopy and Its Determinants among Residents of Ido-Ekiti, Southwestern Nigeria. PLOS ONE. 2023. 18(2):e0280981.

5. Agaba BB, Rugera SP, Mpirirwe R, Atekat M, Okubal S, Masereka K, et al. Asymptomatic Malaria Infection, Associated Factors and Accuracy of Diagnostic Tests in a Historically High Transmission Setting in Northern Uganda. Malaria Journal. 2022. 21(1):392.

6. Wångdahl A, Bogale RT, Eliasson I, Broumou I, Faroogh F, Lind F, et al. Malaria Parasite Prevalence in Sub-Saharan African Migrants Screened in Sweden: A Cross-Sectional Study. The Lancet Regional Health – Europe. 2023. 27.

7. Beavogui AH, Delamou A, Camara BS, Camara D, Kourouma K, Camara R, et al. Prevalence of Malaria and Factors Associated with Infection in Children Aged 6 months to 9 years in Guinea: Results from a National Cross-Sectional Study. Parasite Epidemiol Control. 2020. 11:e00162.

8. Cherif MS, Dahal P, Beavogui AH, Delamou A, Lama EK, Camara A, et al. Malaria Epidemiology and Anti-Malarial Drug Efficacy in Guinea: A Review of Clinical and Molecular Studies. Malar J. 2021. 20(1):272.

9. Institut National de la Statistique. Enquête Démographique et de Santé et à Indicateurs Multiples (Eds-Mics). Conakry: Institut National de la Statistique; 2013.

10. Carnevale P, Toto JC, Guibert P, Keita M, Manguin S. Entomological Survey and Report of a Knockdown Resistance Mutation in the Malaria Vector Anopheles Gambiae from the Republic of Guinea. Trans R Soc Trop Med Hyg. 2010. 104(7):484–9.

11. Programme National de Lutte Contre le Paludisme. Plan StratéGique National De Lutte Contre Le Paludisme 2013 - 2017. Conakry: Programme National de Lutte Contre le Paludisme; 2014.

12. République de Guinée. EnquêTe Sur Les Indicateurs Du Paludisme Et De L’anéMie En GuinéE (Eipag). Conakry: République de Guinée; 2021.

